# Adherence to the Eatwell Guide and cardiometabolic, cognitive and neuroimaging parameters: An analysis from the PREVENT Dementia study

**DOI:** 10.1101/2023.05.09.23289734

**Authors:** Sarah Gregory, Alex Griffiths, Amy Jennings, Fiona C Malcomson, Jamie Matu, Anne-Marie Minihane, Graciela Muniz-Terrera, Craig W. Ritchie, Solange Parra-Soto, Emma Stevenson, Rebecca Townsend, Nicola Ward, Oliver Shannon

**Affiliations:** Edinburgh Dementia Prevention, Centre for Clinical Brain Sciences, University of Edinburgh, Edinburgh, UK; School of Health, Leeds Beckett University, Leeds, UK; Norwich Medical School, University of East Anglia, Norwich, UK; Centre for Public Health, Institute for Global Food Security, Queen’s University Belfast, Belfast, UK; Human Nutrition & Exercise Research Centre, Population Health Sciences Institute, Faculty of Medicine Sciences, Newcastle University, Newcastle upon Tyne, UK; Ohio University Heritage College of Osteopathic Medicine, Ohio University, Ohio, USA; Scottish Brain Sciences, Edinburgh, UK; Department of Nutrition and Public Health, Universidad del Bío-Bío, Chillan 3780000, Chile; School of Cardiometabolic and Metabolic Health, University of Glasgow, UK; School of Biomedical, Nutritional and Sport Sciences, Newcastle University, Newcastle upon Tyne, UK

**Keywords:** diet, Eatwell Guide, cohort study, cardiometabolic health

## Abstract

**Background:** The Eatwell guide reflects the UK government’s recommendations for a healthy and balanced diet. Previous research has identified associations between healthy eating patterns and both cardiovascular and brain health, although there is little evidence specifically focusing on the Eatwell Guide. To date no research has investigated associations between the Eatwell Guide and risk for future dementia.

**Methods:** Data from the PREVENT dementia cohort study baseline visit was used in this analysis. Binary and graded Eatwell Guide scores (BEWG, GEWG) were created from a self-reported Food Frequency Questionnaire. The CAIDE score was included as the primary outcome measure to represent risk for future Alzheimer’s disease. Secondary outcome measures included cardiometabolic health measures and brain health measures. Generalised additive models were run in R.

**Results:** A total of 517 participants were included in the analysis, with a mean BEWG score of 4.39 (± 1.66) (out of a possible 12 points) and GEWG score of 39.88 (± 6.19) (out of a possible 60 points). There was no significant association between either Eatwell Guide score and the CAIDE score (BEWG β: 0.07, 95% confidence interval (CI): −0.07, 0.22; GEWG β: 0.02, 95% CI: - 0.02, 0.06) or any measures of brain health. There was a significant association between higher GEWG score and lower systolic and diastolic blood pressure and body mass index (BMI) (systolic β: −0.24, 95% CI: −0.45, −0.03; diastolic β: −0.16, 95% CI: −0.29, −0.03; BMI β: −0.09, 95% CI: −0.16, −0.01).

**Conclusions:** Although not directly associated with the CAIDE score, the Eatwell Guide dietary pattern may be beneficial for dementia prevention efforts through the modification of hypertension and obesity, which are both known risk factors for dementia. Future work could replicate these findings in other UK-based cohorts as well as further development of Eatwell Guide scoring methodologies.

## Introduction

Healthy eating behaviours have been associated with reduced risk of all-cause mortality and many chronic age-related conditions (1, 2). Indeed, one in seven UK deaths and one in five premature deaths in the UK is thought to be attributable to poor diet (3). The role of healthier dietary behaviours is critical in the context of a globally aging population, particularly for cardiometabolic and brain health, with dementia and heart disease the leading causes of death in the UK (4).

The ‘Eatwell Guide’ (EWG) communicates the UK government recommendations for a healthy and balanced diet (5). The EWG promotes the consumption of fruit and vegetables, oily and other fish, total fibre, sustainable protein sources, wholegrains and fibre-rich carbohydrate sources, and adequate fluid intake, whilst limiting consumption of red and processed meat, total salt, free sugars, saturated fatty acids (SFAs) and total fat (5, 6).

Few studies have explored associations between overall adherence to the EWG and health. In one cross-cohort analysis of data from EPIC-Oxford, One Million Women study and UK Biobank aiming to understand health impacts and environmental footprints of the EWG, higher adherence to the EWG was associated with a reduced risk of mortality (7). This study by Scheelbeck *et al* is the first to create an empirical score from the EWG and investigate associations with health outcomes, applying a binary scoring method for nine of the EWG groups. Analyses applying the same EWG scoring methodology in post-menopausal women in the UK Women’s Cohort Study (UKWCS) reported that higher adherence to the EWG was associated with lower weight, waist circumference and body mass index (BMI) (8). Further, greater adherence to the EWG at baseline was associated with smaller increases in waist circumference and lower risk of abdominal obesity over 4 years (8). Whilst these provisional findings are promising, they are restricted to a limited number of health outcomes. Moreover, Scheelbeek *et al* did not incorporate all EWG components into their score due to availability of data across datasets and scored each EWG component on a binary basis (i.e., points awarded for achieving a dietary goal), which may fail to capture more nuanced differences in diet quality between individuals (e.g., by partially meeting an EWG recommendation).

The EWG has a number of notable similarities to a Mediterranean dietary pattern (MedDiet). An analysis of adherence to a MedDiet in the PREVENT dementia cohort, a UK and Ireland midlife cohort used in this current analysis, showed significant associations between higher adherence to the MedDiet and lower systolic and diastolic blood pressure and BMI, particularly in female participants (9). Given the conceptual overlap between the EWG and the MedDiet, it is reasonable to explore associations between the EWG and cardiovascular and brain health.

Associations between EWG adherence and risk of dementia are currently unknown. Indeed, the SACN (2018) review on *Diet, Cognitive Impairment and Dementia* identified a gap in the research about UK healthy eating recommendations and dementia risk (10). The aim of this current study was to develop a new binary and graded scoring methodologies for EWG adherence, building on the initial methodology developed by Scheelbeek *et al*, in order to explore associations between adherence to the EWG and risk for dementia, cardiometabolic and brain health in a cohort of midlife adults in the UK and Ireland. Furthermore, a comparison was made between the EWG scores and MedDiet scores within the cohort, to explore the comparability of these two models to capture healthy eating.

## Methods

### PREVENT Dementia Programme

The data used in this study is drawn from the baseline visit of the PREVENT dementia programme (PREVENT) (11–13). PREVENT is a prospective cohort study of 700 participants aged 40 to 59 years of age at baseline, at least half of whom have a parental history of dementia, were fluent in English, and who were free of dementia at study entry. Participants were only excluded if they were unable to tolerate the study protocol, including any contraindications for brain magnetic resonance imaging (MRI). Participants were recruited from five centres in the UK and Ireland (Cambridge, Dublin, Edinburgh, London, and Oxford) through memory clinics when in attendance as the family member of a patient, advertisements, research registries and word of mouth. Participants completed physical health and cognitive assessments at the baseline visit as well as providing information on risk factors for future neurodegeneration through a series of self-report questionnaires.

### Ethical Approval and Consent to Participate

The study was conducted according to the guidelines laid down in the Declaration of Helsinki and all procedures involving human participants were approved by the London-Camberwell St Giles National Health Service Research Ethics Committee (REC reference 12/LO/1023). Written informed consent was provided by all participants prior to any protocol procedures.

### Calculation of Eatwell Guide scores

Dietary data were collected with the Scottish Collaborative Group Food Frequency Questionnaire (SCG-FFQ) (14, 15). The SCG-FFQ was self-administered by each participant. It begins with clear instructions on the first page of the questionnaire including pictures of portion sizes, which all participants were instructed to read before self-reporting their diet. Study staff were on hand to answer any questions that participants had and to check for missing data prior to the participant leaving the site. The SCG-FFQ has been validated in several populations in the UK as a self-report tool (14, 15). Compared to a 7-day food diary, there were moderate correlations with SCQ-FFQ derived nutrients for energy (kJ, r_s_: 0.37, p<0.001), percentage energy from fat (r_s_: 0.53, p<0.001), percentage energy from SFA (r_s_: 0.55, p<0.001), percentage energy from protein (r_s_: 0.55, p<0.001), percentage energy from carbohydrates (r_s_: 0.69, p<0.001) and total sugars (r_s_: 0.62, p<0.001) (14). A second study assessing validity in older adults (aged 65 and above) in Scotland compared the SCG-FFQ to a four-day weighted food record, Spearman rank correlation was greater than 0.2 for all nutrients of interest for EWG score calculation with the exception of fat (15). The SCQ-FFQ asks participants to report their consumption of 175 foods and drinks over the previous two to three months. The SCG-FFQ was completed at the baseline visit, with repeated dietary data collection currently ongoing in follow-up visits (Visit 2; 2-4 years post-baseline; Visit 3: 5-8 years post baseline). A comprehensive nutritional breakdown is available for each participant in addition to food level responses. Daily nutrient intake was calculated from the food intake data using the McCance and Widdowson 2021 dataset (16). Intakes of carbohydrates, proteins, total fats and SFA were converted into calorie values to calculate the percentage of calories from each food group included in the diet.

Two EWG scores were created, one applying a binary scoring methodology, and one a graded score (hereafter referred to as the binary EWG and graded EWG, respectively). Full details of scoring methodologies are available in the supplementary materials (Supplementary Table S1). Each score awarded points for adherence to EWG criteria for the following food and nutrient groups; carbohydrates, proteins, fats, SFA, fibre, sugars, salt, total kilocalories, fruit and vegetables, fish, red and processed meats, and water. For the binary scoring, the method was modelled on a traditional Mediterranean Diet Adherence Screener (MEDAS) score (17); participants were awarded 1 point if they met criteria for the nutritional or food component, else 0 points were awarded, with a total possible score of 12. Intake values were not rounded up for any of the components. For example, a participant would be awarded one point if ≥50% of calories reported in the diet were from carbohydrates and 0 points for <50% of calories from carbohydrates. The graded score was modelled on the Panagiotakos Pyramid MedDiet score (18), with 0 to 5 points allocated according to level of compliance with the EWG recommendations, with a total possible score of 60. 5 points was awarded if a participant met the EWG recommendations for a food or macronutrient group. 0 points were awarded for achieving less than half of the recommended intake for healthy foods (carbohydrates, proteins, fibre, fruit and vegetables, fish and water) and for consuming 1.5 times the recommended limit for unhealthy foods (fats, SFA, sugars, salts, red and processed meats). Taking carbohydrates as an example for the binary EWG score 1 point was awarded if ≥50% of calories reported in the diet were from carbohydrates and 0 points for <50% of calories from carbohydrates; for the graded EWG score 5 points were awarded for ≥50% of calories reported in the diet were from carbohydrates, 4 points for ≥43.75% and <50% of calories from carbohydrates, 3 points for ≥37.5% and <43.75% of calories from carbohydrates, 2 points for ≥31.25% and <37.5% of calories from carbohydrates, 1 point for ≥25% and <31.5% of calories from carbohydrates and 0 points for <25% of calories from carbohydrates.

### Calculation of Mediterranean diet scores

Three MedDiet scores (the MEDAS score, the MEDAS continuous and the MedDiet Pyramid (Pyramid) score) were calculated using previously published scoring methods. Briefly, the MEDAS score was calculated using a binary scoring method, whereby participants were allocated 0 or 1 points for each of 14 food groups depending on whether they met consumption criteria (19). The MEDAS continuous was developed by Shannon *et al* with points allocated for the same consumption criteria as MEDAS but on a continuous scale from 0 to 1, depending upon proximity to the dietary target, as opposed to binary allocations (20). Similarly, the Pyramid score was also coded on a continuous scale of 0 to 1 with a total possible score of 15 points (21). Continuous scores have been shown to have more sensitivity to detecting differences in diet quality, particularly in a UK population, where they have shown stronger associations with better cognition (20) and reduced dementia risk (22) compared with binary scores.

### CAIDE score

The Cardiovascular Risk Factors, Ageing and Dementia (CAIDE) risk score, an estimate of 20-year dementia risk based on risk factors in midlife, was calculated for all participants. The CAIDE score was originally developed in the FINGER study, and ranges from 0 to 18 points with higher scores representing greater dementia risk (23). The CAIDE score combines a number of non-modifiable and potentially modifiable risk factors. It was selected as it has previously been associated with a number of neuroimaging outcomes in the PREVENT dementia cohort (24–28). Additionally, the CAIDE score has been suggested as an appropriate surrogate outcome measure in lifestyle-based multidomain prevention trials (29), and the FINGER multi-domain intervention (which included changes to diet, as well as exercise, cognitive training and management of metabolic and vascular risk factors) significantly reduced the CAIDE score (30, 31), demonstrating the score has the potential to be responsive to lifestyle factors. The CAIDE score was calculated using self-reported age, education and sex, systolic blood pressure (SBP) (mean of triplicate blood pressure readings in supine or seated position recorded at baseline visit), BMI (height and weight recorded at baseline visit, used for BMI calculation), fasting plasma total cholesterol, (analysed in local laboratories at the baseline visit), physical activity (self-reported non-validated questionnaire asking participants how often they complete light, moderate and vigorous exercise; 0 points awarded for never up to 5 points for daily, scores summed across all three categories with higher points reflecting more physical activity) and *APOEε4* carrier status (DNA analysed from blood collected at baseline). The score weighting is presented in Table S2

### Cardiometabolic outcome variables

Data on blood pressure (systolic and diastolic (SBP, DBP)), BMI, and waist-to-hip ratio (WHR)) values (recorded at baseline visit) were extracted from the database. Each of these cardiometabolic measures were collected by trained study staff at the baseline visit. A Framingham Risk Score (FRS) was calculated for each participant using the ‘CVrisk’ package in R (32) and a QRisk3 score was calculated using the ‘QRISK3’ R package (33). The variables used to create these cardiovascular risk scores are detailed in Supplementary Table S3.

### Cognitive outcome measures

For the purposes of this analysis, the score for the Four Mountains Task (4MT) was selected as the primary cognitive outcome. The 4MT is a novel tablet-administered task designed to assess allocentric processing. Participants are shown an image of four mountains for approximately 10 seconds and after a short interval (∼ 1 second) asked to select which scene they were previously shown from a choice of four image options (34). A total score is derived from 15 trials, with higher marks indicating better performance. This cognitive task remains a research rather than clinical tool and no normative values are currently available. The 4MT has been shown to be sensitive to early neurodegenerative disease (35), has previously been associated with CAIDE scores in the PREVENT cohort (36), and has also previously been associated with the MedDiet in a European cohort study (37).

### Magnetic resonance imaging (MRI) variables

MRI scans were collected using 3T Siemens scanners (Verio, PRISMA, Prisma Fit, Skyra), with data for this study extracted from T1-weighted structural scans processed with FreeSurfer (version 7.1.0) and FLAIR MRI using SPM8. Further details of imaging acquisition, processing and analysis are available in the cohort baseline data descriptive paper (13). Derived variables were extracted from the dataset to include left and right hippocampal volume, left and right hippocampal thickness, white matter hyperintensity volume (cube-transformed and corrected for total intracranial volume) and total estimated intra-cranial volume. Further details on the imaging acquisition and processing in the PREVENT dataset can be found elsewhere (38).

### Perception of healthy eating

Participants were asked to indicate (yes or no) if they felt they ate a healthy diet. No further context was provided as to what defined a healthy diet and there was no set time period, rather participants were asked to respond about how they felt generally about their diet. Self-reported diet quality was included to investigate if participants’ beliefs about their healthy eating habits aligned with higher EWG guide scores as a model of healthy eating.

### Covariates

Several covariates were assessed, including age, sex, years of education, *APOE⍰4*, parental history of dementia (self-reported), socioeconomic status (SES) group and physical activity. SES group was determined according to self-reported occupation using the National Statistics socio-economic classification (NS-SEC: https://www.ons.gov.uk/methodology/classificationsandstandards/otherclassifications/thenationalstatisticssocioeconomicclassificationnssecrebasedonsoc2010) and grouped in low, middle and high socioeconomic group or into a not in employment group. The not in employment group included both participants who reported they were unemployed and those who had taken early retirement. As total kilocalories were included in the EWG scores, the analyses were not adjusted for total energy intake. For analysis including the CAIDE score as the outcome measure, only parental history of dementia and physical activity were included as covariates so as not to over-correct the model. Where the FRS or QRisk3 was the outcome variable of interest, years of education *APOE*⍰*4*, parental history of dementia and physical activity were included as covariates. The National Adult Reading Test (NART) score was included as an additional covariate in the 4MT analysis as a measure of premorbid intelligence. Finally, for all the brain imaging models, intracranial volume was included as a covariate.

### Statistical Analysis

All statistical analyses were completed using R (Version 4.1.0). Descriptive statistics were calculated for all participants. Where necessary, to ensure the fulfilment of distributional assumptions of the models fitted, data was transformed. For the main analysis, we excluded participants with missing data in the exposure, outcome, and covariate variables of interest from the analysis (n=183 missing, data missing at random; missing dietary data or implausible calorie intake (n=106), missing outcome data (n=77); additionally a further 38 participants were excluded from analyses including neuroimaging outcome measures due to missing MRI data or incidental MRI findings. Relationships between the binary EWG (BEWG) and graded EWG (GEWG) scores and the MedDiet scores were assessed using correlations. As the BEWG and GEWG scores were slightly skewed, generalised additive models were run. First, we tested the cohort as a whole and fitted univariate and fully adjusted generalised additive models to test for associations between BEWG and GEWG scores and the CAIDE score. The fully adjusted model included parental history of dementia, physical activity scores and SES group. We then ran univariate and fully adjusted generalised additive models to test for associations between BEWG and GEWG scores and measures of cardiometabolic health (SBP, DBP, BMI, WHR, FRS, QRisk3) and brain health (4MT total score, cube-transformed white matter lesion volume, left and right hippocampal volume, and left and right hippocampal thickness). Generalised linear models were used to assess associations between BEWG and GEWG scores and self-rated diet quality. Covariates included in each model are detailed in the tables of results. Finally, an exploratory component level analysis was run for the CAIDE score (as the primary outcome) and for all other outcomes with a statistically significant fully adjusted model. A further exploratory analysis tested for any differences in outcomes with the GEWG score by SES group. Finally, the BEWG was split around the median (4) to categorise participants to lower or higher adherence. Logistic regression models were run to test for associations between categorisation around the median and the primary and secondary outcome measures. Primary and secondary analyses were adjusted for multiple comparisons using the Benjamini-Hochberg False Discovery Rate (FDR) procedure. A formal sample size calculation was not undertaken as this was a secondary analysis of a large observational study.

## Results

### Descriptive statistics

A total of 517 participants were included in the primary analyses which investigated CAIDE risk scores and cardiometabolic health. Additional analyses involved fewer participants due to missing data, with sample sizes for each outcome detailed in Table 1. The sample included more women (59.6%), had a similar number of participants with and without a parental history of dementia (52.8% vs 47.2%), with 38.3% *APOE⍰4* carriers. Most participants fell in the highest SES group according to their occupations (64.6%), with a high number of years of education reported in the sample (16.72 (SD 3.31) years). See Table 1 for full demographic and descriptive details.

**Table 1:**
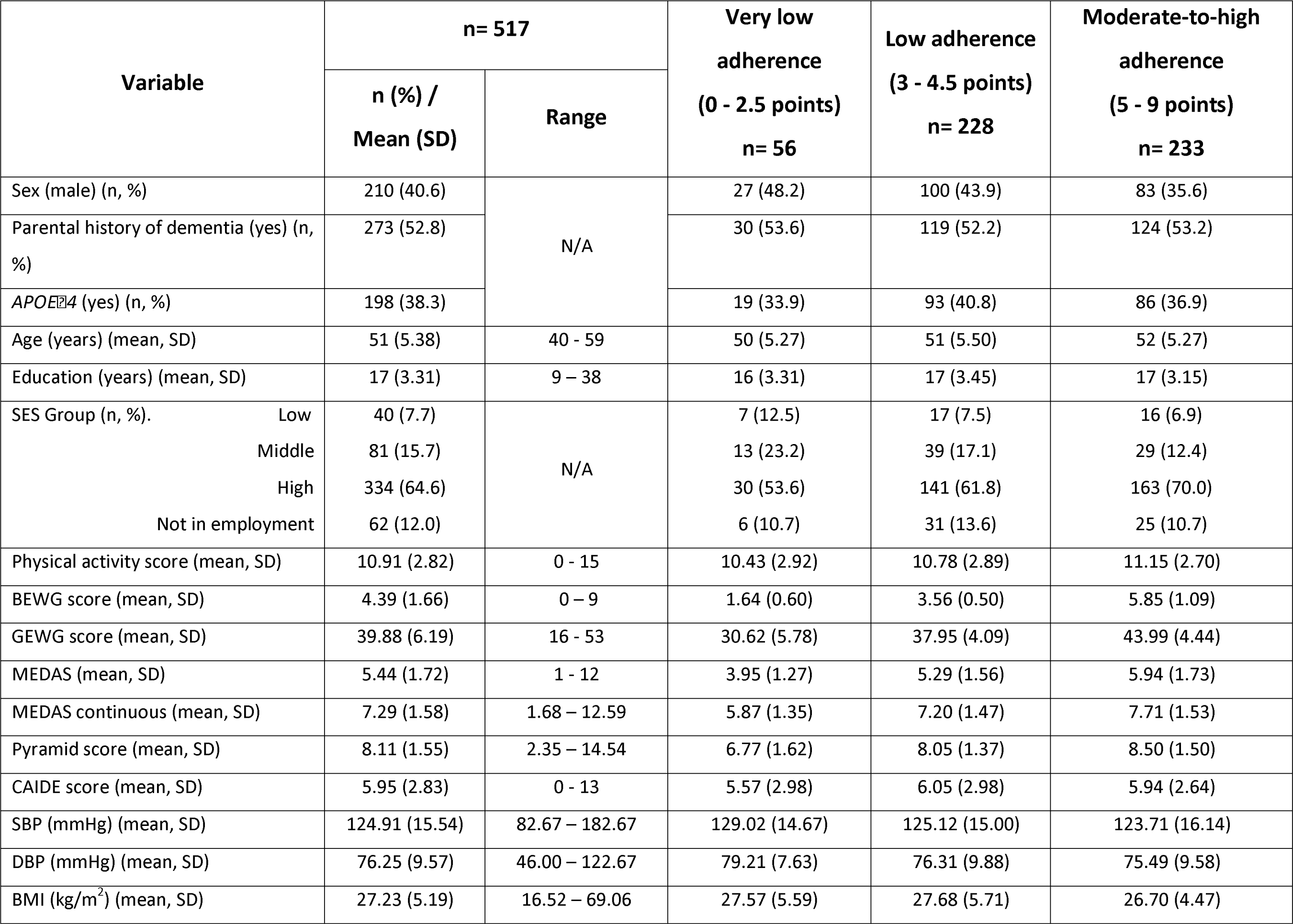
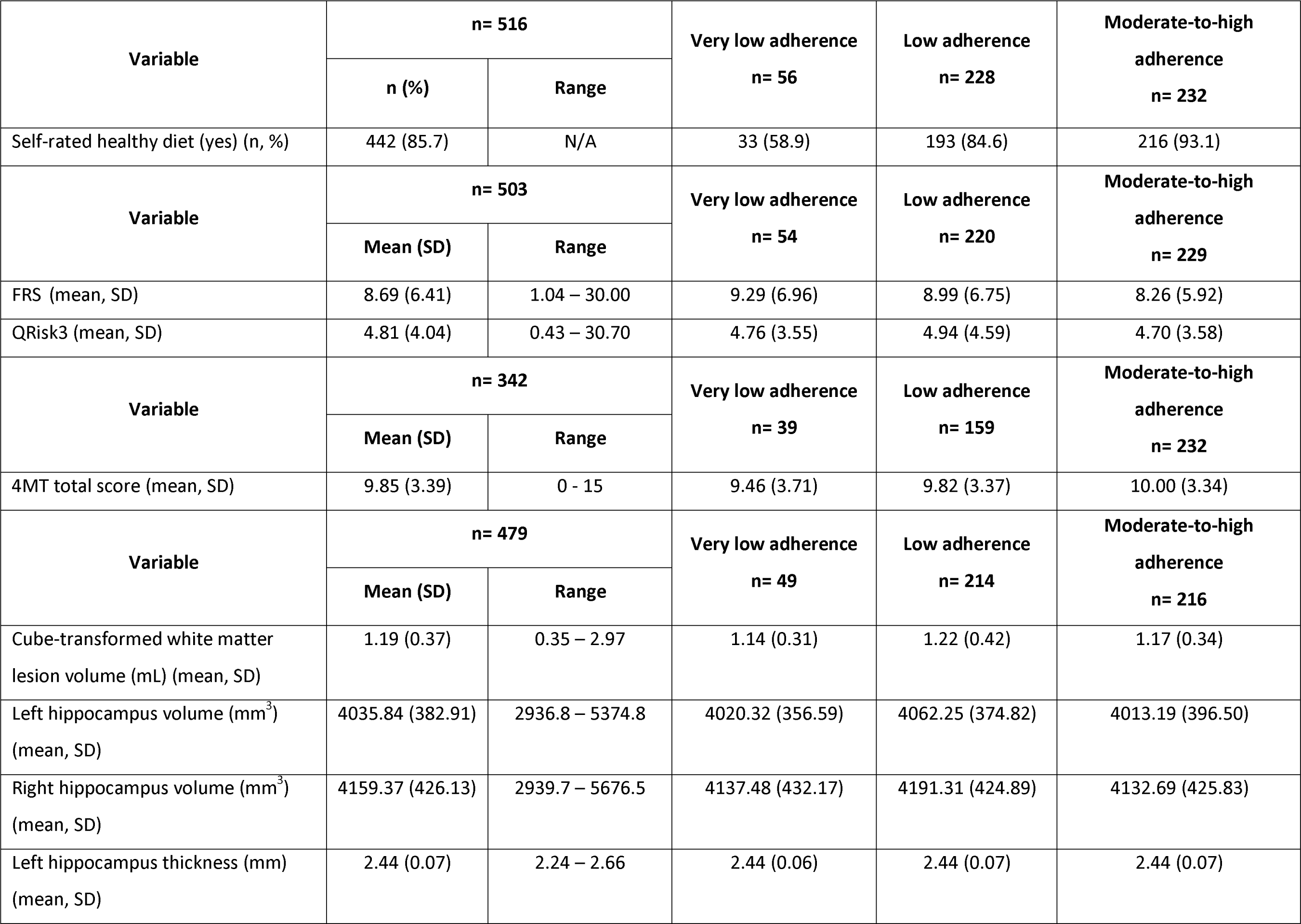
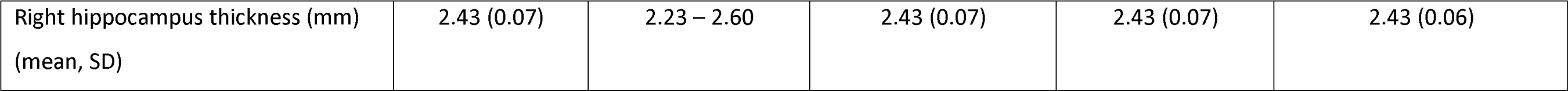
Demographic and descriptive statistics of sample included in Eatwell Guide score analysis. 4MT: Four Mountains Test; BMI: body mass index; DBP: diastolic blood pressure; EWG: Eatwell Guide; FRS: Framingham Risk Score; SBP: systolic blood pressure; SES: socioeconomic status.

| Variable | n= 517 |  | Very low adherence<br>(0 - 2.5 points)<br>n= 56 | Low adherence<br>(3 - 4.5 points)<br>n= 228 | Moderate-to-high adherence<br>(5 - 9 points)<br>n= 233 |
| --- | --- | --- | --- | --- | --- |
|  | n (%) / Mean (SD) | Range |  |  |  |
| Sex (male) (n, %) | 210 (40.6) | N/A | 27 (48.2) | 100 (43.9) | 83 (35.6) |
| Parental history of dementia (yes) (n, %) | 273 (52.8) |  | 30 (53.6) | 119 (52.2) | 124 (53.2) |
| APOEε4 (yes) (n, %) | 198 (38.3) | | 19 (33.9) | 93 (40.8) | 86 (36.9) |
| Age (years) (mean, SD) | 51 (5.38) | 40 - 59 | 50 (5.27) | 51 (5.50) | 52 (5.27) |
| Education (years) (mean, SD) | 17 (3.31) | 9 – 38 | 16 (3.31) | 17 (3.45) | 17 (3.15) |
| SES Group (n, %). |  | N/A |  |  |  |
| Low | 40 (7.7) |  | 7 (12.5) | 17 (7.5) | 16 (6.9) |
| Middle | 81 (15.7) |  | 13 (23.2) | 39 (17.1) | 29 (12.4) |
| High | 334 (64.6) |  | 30 (53.6) | 141 (61.8) | 163 (70.0) |
| Not in employment | 62 (12.0) |  | 6 (10.7) | 31 (13.6) | 25 (10.7) |
| Physical activity score (mean, SD) | 10.91 (2.82) | 0 - 15 | 10.43 (2.92) | 10.78 (2.89) | 11.15 (2.70) |
| BEWG score (mean, SD) | 4.39 (1.66) | 0 – 9 | 1.64 (0.60) | 3.56 (0.50) | 5.85 (1.09) |
| GEWG score (mean, SD) | 39.88 (6.19) | 16 - 53 | 30.62 (5.78) | 37.95 (4.09) | 43.99 (4.44) |
| MEDAS (mean, SD) | 5.44 (1.72) | 1 - 12 | 3.95 (1.27) | 5.29 (1.56) | 5.94 (1.73) |
| MEDAS continuous (mean, SD) | 7.29 (1.58) | 1.68 – 12.59 | 5.87 (1.35) | 7.20 (1.47) | 7.71 (1.53) |
| Pyramid score (mean, SD) | 8.11 (1.55) | 2.35 – 14.54 | 6.77 (1.62) | 8.05 (1.37) | 8.50 (1.50) |
| CAIDE score (mean, SD) | 5.95 (2.83) | 0 - 13 | 5.57 (2.98) | 6.05 (2.98) | 5.94 (2.64) |
| SBP (mmHg) (mean, SD) | 124.91 (15.54) | 82.67 – 182.67 | 129.02 (14.67) | 125.12 (15.00) | 123.71 (16.14) |
| DBP (mmHg) (mean, SD) | 76.25 (9.57) | 46.00 – 122.67 | 79.21 (7.63) | 76.31 (9.88) | 75.49 (9.58) |
| BMI (kg/m<sup>2</sup>) (mean, SD) | 27.23 (5.19) | 16.52 – 69.06 | 27.57 (5.59) | 27.68 (5.71) | 26.70 (4.47) |

| Variable | n= 516 |  | Very low adherence<br>n= 56 | Low adherence<br>n= 228 | Moderate-to-high<br>adherence<br>n= 232 |
| --- | --- | --- | --- | --- | --- |
|  | n (%) | Range |  |  |  |
| Self-rated healthy diet (yes) (n, %) | 442 (85.7) | N/A | 33 (58.9) | 193 (84.6) | 216 (93.1) |
| Variable | n= 503 |  | Very low adherence<br>n= 54 | Low adherence<br>n= 220 | Moderate-to-high<br>adherence<br>n= 229 |
|  | Mean (SD) | Range |  |  |  |
| FRS (mean, SD) | 8.69 (6.41) | 1.04 – 30.00 | 9.29 (6.96) | 8.99 (6.75) | 8.26 (5.92) |
| QRisk3 (mean, SD) | 4.81 (4.04) | 0.43 – 30.70 | 4.76 (3.55) | 4.94 (4.59) | 4.70 (3.58) |
| Variable | n= 342 |  | Very low adherence<br>n= 39 | Low adherence<br>n= 159 | Moderate-to-high<br>adherence<br>n= 232 |
|  | Mean (SD) | Range |  |  |  |
| 4MT total score (mean, SD) | 9.85 (3.39) | 0 - 15 | 9.46 (3.71) | 9.82 (3.37) | 10.00 (3.34) |
| Variable | n= 479 |  | Very low adherence<br>n= 49 | Low adherence<br>n= 214 | Moderate-to-high<br>adherence<br>n= 216 |
|  | Mean (SD) | Range |  |  |  |
| Cube-transformed white matter lesion volume (mL) (mean, SD) | 1.19 (0.37) | 0.35 – 2.97 | 1.14 (0.31) | 1.22 (0.42) | 1.17 (0.34) |
| Left hippocampus volume (mm<sup>3</sup>) (mean, SD) | 4035.84 (382.91) | 2936.8 – 5374.8 | 4020.32 (356.59) | 4062.25 (374.82) | 4013.19 (396.50) |
| Right hippocampus volume (mm<sup>3</sup>) (mean, SD) | 4159.37 (426.13) | 2939.7 – 5676.5 | 4137.48 (432.17) | 4191.31 (424.89) | 4132.69 (425.83) |
| Left hippocampus thickness (mm) (mean, SD) | 2.44 (0.07) | 2.24 – 2.66 | 2.44 (0.06) | 2.44 (0.07) | 2.44 (0.07) |
| Right hippocampus thickness (mm)<br>(mean, SD) | 2.43 (0.07) | 2.23 – 2.60 | 2.43 (0.07) | 2.43 (0.07) | 2.43 (0.06) |

### Dietary score descriptive statistics

The sample had a mean BEWG score of 4.39 (standard deviation (SD) 1.66) (range 0 to 9) and a mean GEWG score of 39.88 (SD 6.19) (range 16 to 53). Women had higher BEWG scores compared to men (4.55 (SD 1.66) vs 4.15 (SD 1.65), t: 2.70, p: 0.007) however this difference was smaller when comparing women to men for the GEWG scores (40.31 (SD 6.16) vs 39.24 (SD 6.20), t: 1.94, p: 0.05). Participants in the low SES group had lower GEWG scores than the high SES group (Low: 37.50 (SD 6.57); High: 40.55 (SD 5.90), p: 0.003), with no significant difference in BEWG scores by SES group. Age was significantly associated with higher BEWG and GEWG scores (BEWG β: 0.03, standard error (SE): 0.01, 95% confidence interval (CI): 0.002, 0.06, p: 0.03; GEWG β: 0.14, SE: 0.05, 95% CI: 0.04, 0.24, p: 0.004). Higher physical activity scores were associated with higher GEWG, but not BEWG, scores (β: 0.22, SE: 0.10, 95% CI: 0.03, 0.41, p: 0.02). There were no differences in BEWG or GEWG score by parental history of dementia or *APOE*⍰*4* status. There was no significant association between the total number of years of education and either the BEWG or GEWG scores. A breakdown of the number of contributing component information for each score is presented in Supplementary Table S4. All participants were consuming more than 5% of calories from sugars and so no participants were awarded a point for this component using the BEWG scoring methodology. This may be due to the way sugars were calculated from the SCQ-FFQ and is explored further in the discussion.

### Correlations between EWG and MedDiet scores

The BEWG and GEWG scores were highly correlated with each other (R: 0.77, p<0.001). BEWG and GEWG scores were correlated with MedDiet scores to explore the similarity between the dietary patterns. All scores were moderately correlated (r= 0.3-0.4), with moderate correlations between the BEWG and GEWG scores with the three MedDiet scores (MEDAS, MEDAS continuous, Pyramid) (see Figure 1).

### Analytical statistics

#### CAIDE

There was no significant association between the BEWG score or GEWG score and the CAIDE score in unadjusted or fully adjusted models (Fully adjusted scores; BEWG β: 0.07, 95% CI: - 0.07, 0.22; GEWG β: 0.02, 95% CI: −0.02, 0.06) (see Table 2). Meeting fat requirements (i.e. ≤35% calories from fat) for both the BEWG and the GEWG scores was associated with a higher CAIDE score (Fully adjusted scores; BEWG β: 0.61, 95% CI: 0.12, 1.11; GEWG β: 0.24, 95% CI: 0.01, 0.48), with no other associations seen at the food or nutritional component level, or when participants were categorised into high and low adherence by the median (see Supplementary Table S5, S6, S10 and S11).

**Table 2:** Table of generalised additive models for associations between EWG and EWG graded with CAIDE and cardiometabolic health outcomes. BMI: body mass index; CI: confidence interval; EWG: Eatwell Guide score; FRS: Framingham Risk Score; WHR: waist-to-hip ratio. CAIDE score fully adjusted model includes parental history of dementia, physical activity score and socioeconomic status as covariates. Systolic/diastolic blood pressure, BMI and WHR fully adjusted models include age, sex, education, *APOEε4*, parental history of dementia, physical activity score and socioeconomic status as covariates. FRS and QRisk3 fully adjusted models include education, *APOEε4*, parental history of dementia, physical activity score and socioeconomic status as covariates. p<0.05 after False Discovery Rate (FDR) adjustment.

| Dietary score | Unadjusted |  |  |  | Fully adjusted |  |  |  |
| --- | --- | --- | --- | --- | --- | --- | --- | --- |
| | $\beta$ | SE | 95% CI | p | $\beta$ | SE | 95% CI | p |
| **CAIDE** |  |  |  |  |  |  |  |  |
| **EWG** | 0.03 | 0.07 | -0.11, 0.18 | 0.64 | 0.07 | 0.07 | -0.07, 0.22 | 0.32 |
| **EWG graded** | 0.0001 | 0.02 | -0.04, 0.04 | 0.96 | 0.02 | 0.02 | -0.02, 0.06 | 0.36 |
| **Systolic Blood Pressure** |  |  |  |  |  |  |  |  |
| **EWG** | -0.77 | 0.41 | -1.59, 0.05 | 0.06 | -0.45 | 0.39 | -1.23, 0.33 | 0.25 |
| **EWG graded** | -0.27 | 0.11 | -0.49, -0.05 | 0.01 | -0.24 | 0.11 | -0.45, -0.03 | 0.03 |
| **Diastolic Blood Pressure** |  |  |  |  |  |  |  |  |
| **EWG** | -0.44 | 0.25 | -0.94, 0.07 | 0.08 | -0.22 | 0.24 | -0.70, 0.27 | 0.37 |
| **EWG graded** | -0.19 | 0.07 | -0.32, -0.05 | 0.006 | -0.16 | 0.06 | -0.29, -0.03 | 0.01 |
| **BMI** |  |  |  |  |  |  |  |  |
| **EWG** | -0.14 | 0.14 | -0.41, 0.14 | 0.32 | -0.04 | 0.14 | -0.31, 0.23 | 0.78 |
| **EWG graded** | -0.12 | 0.04 | -0.19, -0.05 | 0.001<sup>†</sup> | -0.09 | 0.04 | -0.16, -0.01 | 0.02 |
| **WHR** |  |  |  |  |  |  |  |  |
| **EWG** | -0.005 | 0.003 | -0.01, 0.0003 | 0.06 | -0.0006 | 0.002 | -0.005, 0.004 | 0.79 |
| **EWG graded** | -0.002 | 0.0007 | -0.003, -0.0002 | 0.02 | -0.0006 | 0.0006 | -0.002, 0.0007 | 0.32 |
| **FRS** |  |  |  |  |  |  |  |  |
| **EWG** | -0.27 | 0.17 | -0.61, 0.07 | 0.11 | -0.25 | 0.17 | -0.59, 0.09 | 0.14 |
| **EWG graded** | -0.09 | 0.05 | -0.18, 0.002 | 0.05 | -0.06 | 0.05 | -0.16, 0.03 | 0.17 |
| **QRisk3** |  |  |  |  |  |  |  |  |
| **EWG** | -0.08 | 0.11 | -0.30, 0.14 | 0.46 | -0.06 | 0.11 | -0.28, 0.16 | 0.58 |
| **EWG graded** | -0.01 | 0.03 | -0.07, 0.04 | 0.61 | 0.0003 | 0.03 | -0.06, 0.06 | 0.99 |

### Cardiometabolic health

There were no significant associations between BEWG scores and SBP, DBP or BMI, and no significant associations between either scoring methodology and WHR, FRS or QRisk3 scores. In contrast, higher GEWG scores were associated with lower SBP and DBP (fully adjusted SBP β: - 0.24, 95% CI: −0.45, −0.03; DBP β: −0.16, 95% CI: −0.29, −0.03), as well as with lower BMI (β: −0.09, 95% CI: −0.16, −0.01) (see Table 2). None of the fully adjusted models remained significant after FDR adjustment for multiple comparisons. Higher scores awarded for the GEWG total fat component (i.e. eating fewer total calories from fat and therefore being closer to achieving ≤35% calories from fat) were associated with higher SBP (β: 1.53, 95% CI: 0.28, 2.78). Conversely, having higher scores for fibre (indicating being closer to achieving the EWG dietary target of ≥22.6g/d of fibre), fruits and vegetables (indicating being closer to achieving the EWG dietary target of ≥400g/d of fruits and vegetables), and fish (indicating being closer to achieving the EWG dietary target of ≥10g/d of fish) GEWG score components was associated with significantly lower SBP (fibre β: −0.97, 95% CI: −1.69, −0.26; fruits and vegetables β: −1.09, 95% CI: −1.97, −0.20; fish β: −1.03, 95% CI: −1.75, −0.30). Higher scores for the fibre, fruits and vegetables, and red and processed meat (indicating being closer to achieving the EWG dietary target of ≤70g/d of red or processed meat, i.e. higher scores reflect eating less of this food group) GEWG score components were associated with significantly lower DBP (fibre β: −0.61, 95% CI: −1.05, −0.16; fruits and vegetables β: −0.79, 95% CI: −1.34, −0.24; red and processed meats β: −0.77, 95% CI: −1.34, −0.20). Only higher scores for the fruits and vegetables graded EWG score component were associated with lower BMI (β: −0.38, 95% CI: −0.69, −0.07). Low adherence to the EWG (as defined by a score below the median) was associated with higher SBP and DBP. Further details of these associations are provided in Supplementary Tables S7, S8, S9, S10 and S11.

### Four Mountains Test and MRI variables

There was no significant association between either the BEWG or the GEWG score and the 4MT total score (BEWG β: 0.05, 95% CI: −0.18, 0.28; GEWG β: 0.02, 95% CI: −0.04, 0.08) (see Table 3). There were no significant associations between the BEWG or GEWG scores and any MRI variables in the fully adjusted models (see Table 3). In the high SES group only, there was a significant negative association between GEWG scores and left hippocampal volume (see Supplementary Table S9). In the fully adjusted model only, lower adherence to the EWG was associated with higher 4MT total scores, however as this was not statistically significant in the unadjusted model and was conducted as an exploratory analysis this result should be interpreted with caution (see Supplementary Tables S10 and S11).

**Table 3:** Table of generalised additive models for associations between EWG and EWG graded with cardiometabolic risk scores, 4MT score and self-reported healthy eating. CI: confidence interval; 4MT: Four Mountains Test; EWG: Eatwell Guide score; FRS: Framingham Risk Score. 4MT score fully adjusted model includes age, sex, education, *APOEε4*, parental history of dementia, NART score, physical activity score and socioeconomic status as covariates. Self-reported healthy eating score includes age, sex, education, *APOEε4*, parental history of dementia, *APOEε4*, physical activity score and socioeconomic status as covariates. ^¶^p<0.05 after False Discovery Rate (FDR) adjustment.

| Dietary score | Unadjusted |  |  |  | Fully adjusted |  |  |  |
| --- | --- | --- | --- | --- | --- | --- | --- | --- |
| | $\beta$ | SE | 95% CI | p | $\beta$ | SE | 95% CI | p |
| **Four Mountains Test** |  |  |  |  |  |  |  |  |
| **EWG** | 0.04 | 0.11 | -0.18, 0.26 | 0.72 | 0.05 | 0.11 | -0.18, 0.28 | 0.68 |
| **EWG graded** | 0.02 | 0.03 | -0.04, 0.08 | 0.52 | 0.02 | 0.03 | -0.04, 0.08 | 0.58 |
| **White Matter Lesion Volume** |  |  |  |  |  |  |  |  |
| **EWG** | -0.005 | 0.01 | -0.03, 0.02 | 0.64 | -0.005 | 0.01 | -0.02, 0.01 | 0.63 |
| **EWG graded** | -0.001 | 0.003 | -0.007, 0.004 | 0.66 | -0.002 | 0.003 | -0.007, 0.003 | 0.47 |
| **Left Hippocampus** |  |  |  |  |  |  |  |  |
| **EWG** | -17.05 | 8.50 | -34.04, -0.05 | 0.05 | -14.85 | 8.74 | -32.33, 2.63 | 0.09 |
| **EWG graded** | -5.11 | 2.27 | -9.66, -0.57 | 0.02 | -4.28 | 2.38 | -9.04, 0.48 | 0.07 |
| **Right Hippocampus** |  |  |  |  |  |  |  |  |
| **EWG** | -15.43 | 9.50 | -34.44, 3.57 | 0.11 | -13.99 | 9.77 | -33.52, 5.55 | 0.15 |
| **EWG graded** | -1.47 | 2.55 | -6.57, 3.63 | 0.57 | -0.74 | 2.67 | -6.07, 4.60 | 0.78 |
| **Left Hippocampal Thickness** |  |  |  |  |  |  |  |  |
| **EWG** | 0.001 | 0.002 | -0.003, 0.005 | 0.51 | 0.002 | 0.002 | -0.002, 0.006 | 0.40 |
| **EWG graded** | 0.0006 | 0.0005 | -0.0004, 0.002 | 0.24 | 0.0008 | 0.0005 | -0.0003, 0.002 | 0.13 |
| **Right Hippocampal Thickness** |  |  |  |  |  |  |  |  |
| **EWG** | 0.0006 | 0.002 | -0.003, 0.004 | 0.73 | 0.001 | 0.002 | -0.003, 0.005 | 0.54 |
| **EWG graded** | 0.0003 | 0.0005 | -0.0006, 0.001 | 0.49 | 0.0006 | 0.0005 | -0.0004, 0.002 | 0.26 |
| **Self-reported healthy eating** |  |  |  |  |  |  |  |  |
| **EWG** | 0.43 | 0.09 | 0.26, 0.60 | <0.001<sup>†</sup> | 0.38 | 0.09 | 0.21, 0.57 | <0.001<sup>†</sup> |
| **EWG graded** | 0.11 | 0.02 | 0.07, 0.16 | <0.001<sup>†</sup> | 0.10 | 0.02 | 0.05, 0.14 | <0.001<sup>†</sup> |

### Perception of healthy eating

There was a significant association between the positive self-report of eating a healthy diet, and higher BEWG and GEWG scores (BEWG β: 0.92, 95% CI: 0.21, 0.57; GEWG β: 3.71, 95% CI: 0.05, 0.14) (see Table 3). These associations remained significant after FDR adjustment. There was a significant association between the positive self-report of eating a healthy diet and higher BEWG in the middle, high and not in-employment SES groups, but not in the low SES group, and with the GEWG in the high and not in employment SES groups but not low or middle SES groups (see Supplementary Table S9).

## Discussion

Both the BEWG and GEWG scores created in this analysis were moderately correlated with three commonly used MedDiet scores (the MEDAS, MEDAS continuous and Pyramid scores). There were no associations between either EWG score and the primary outcome of the CAIDE score. However, when looking at individual cardiometabolic components of the CAIDE there was an association between higher GEWG scores and lower SBP, DBP and BMI. In particular, achieving more points (indicating being closer to meeting the EWG criteria in full) for fruits and vegetables was associated with better cardiometabolic health. It is important to note that these associations did not remain significant after FDR adjustment and all component level analysis are considered to be exploratory. There were no associations noted between binary or graded EWG scores and brain health as assessed by cognitive or brain volume outcomes. There was a significant association between self-perception of a healthy diet and higher binary and graded EWG scores, with the association strongest in the high SES group as well as in those participants who were not in employment at the time of dietary data collection. This replicates previous research that has shown associations between self-perceived diet and more objective measures of dietary quality (39, 40).

There were no significant associations between either the binary or graded EWG scores and the CAIDE score. The CAIDE score was selected as one of the most commonly used dementia risk scores, with associations between the score and neuroimaging outcomes previously reported in the PREVENT dementia cohort (24–28). Importantly the CAIDE score reflects the accumulation of cardiovascular risk for dementia, factors which may be the most amenable to dietary interventions. However, the CAIDE score is not without limitations and some validation work outside of the original cohort where the score was developed has suggested there is very little discrimination compared to age alone (41), with a limited clinical utility for estimating 10-year dementia risk identified in the UK Biobank cohort (42). Other studies in the USA and The Netherlands conversely have validated the CAIDE score for the prediction of dementia up to four decades later (43) as well as the prediction of cognitive impairment 10 to 15 years later (44, 45), demonstrating the complexity of the ongoing debate about the usefulness of the CAIDE as a predictive dementia risk score. It is also important to consider that many of the components of the CAIDE score would not be modifiable by diet (age, sex, education and *APOEε4*) which may explain the lack of association reported in this analysis. The CAIDE score in the population included in this analysis from the PREVENT cohort is lower compared to the FINGER cohort intervention study where the score was originally developed (PREVENT: 5.95 vs FINGER: 7.76 (intervention) and 7.27 (control) (46)), and it may be that any EWG score associations would only be seen in a cohort with a higher mean CAIDE score where there is more potential for modification. As age is one of the important contributors to the overall CAIDE score, it is worth replicating this analysis between EWG scores and CAIDE score in an older cohort (such as the NICOLA or UK Biobank cohorts (47, 48)) to understand if there is an association in later in midlife, where the mean cohort CAIDE score would be expected to be higher due to age.

Despite no statistically significant associations with the CAIDE score, there were a number of significant associations between the GEWG score and cardiometabolic health which themselves are likely protective of brain health. Importantly these are the elements of the CAIDE score which would be expected to be modifiable by diet. GEWG scores were associated with lower SBP, DBP and BMI. As there were no significant associations between the BEWG score and cardiometabolic health measures, this suggests the GEWG score is more appropriate to apply to this population with partial compliance to EWG criteria important for health. This may reflect previously reported statistics that only 0.1% of the UK population adhere to all nine recommendations (7). In the context of dementia prevention efforts, it is particularly important to note that the GEWG was associated with lower blood pressure and BMI values, given both hypertension and obesity are known midlife risk factors for Alzheimer’s disease (AD) (49). A ten-point change in the GEWG was associated with a 2.4 mmHg reduction in SBP, a 1.6 mmHg reduction in DBP and a 0.9kg/m^2^ reduction in BMI. A 2mmHg reduction in SBP has been estimated to decrease the risk of death from stroke by 10% (50), although larger reductions in SBP may be needed to reduce the risk of dementia with a potential U-shaped association where both low and high BP confers risk (51, 52). Similarly, a 2 mmHg reduction in DBP has been estimated to result in a 17% decrease in hypertension and a 15% reduction of risk from stroke and transient ischaemic attacks (53). In midlife, each 1 unit increase in BMI was associated with a higher risk of dementia in a 38-year follow-up of the Framingham Study (54).

There were no associations seen between either EWG score and any of the brain health outcome measures, with the exception of an association between higher GEWG score and lower left hippocampal volume in the high SES group only. As this was only seen in the left and not right hippocampus and in a single SES group only, it should be interpreted with caution, although previous studies have also found a stronger effect of a healthy diet in the left compared to the right hippocampus (55, 56) and this warrants further research. Exploring whether the EWG scores are associated with functional brain imaging measures as well as with AD pathology (such as amyloid beta, tau and neurofilament light) will also be important next steps for research. The 4MT was selected as the only cognitive measure used in this analysis, and was selected due to previously associations with dementia risk, early neurodegeneration and MedDiet adherence (35–37). Future research should consider whether other measures of cognition included in the PREVENT cohort warrant investigation with dietary quality as measured by the EWG.

Unsurprisingly and reassuringly, there were significant but moderate correlations between the EWG scores and MedDiet scores which demonstrates some overlap in these healthy eating patterns as well as a divergence in how the scores are created. For example, whilst both dietary patterns prioritise the consumption of fruits and vegetables and fish, with limited red and processed meats, the EWG otherwise focuses on a recommended macronutrient intake whilst the MedDiet recommends foods such as olive oil, legumes, and nuts. This should be a noted limitation of the EWG compared to MedDiet scores when translating to public health approaches, as the EWG requires people to know their nutrient intake and understand percentages of intake by calories. Further development of the EWG to translate the recommendations to a more food-based approach, as in the MedDiet and recommended by the Nutrition for Dementia Prevention Working Group (57), will be important. In particular, evidence suggests that using olive oil as the predominant fat in a diet has promise for mitigating vascular risk factors for AD (58).

Meeting, or approaching the set criteria, for fruit and vegetable consumption (≥400g/d) was associated with lower SBP, DBP and BMI. A one-point change on this criteria (indicating being closer to consuming ≥400g/d of fruit and vegetables) was associated with a 1.09 mmHg reduction in SBP, a −0.79 mmHg reduction in DBP, and a −0.38 kg/m^2^ reduction in BMI. This has been seen in a number of studies (59, 60), included in Scheelbeck *et al* where fruit and vegetable consumption was associated with the largest reduction in mortality risk (7). Given adopting dietary change is complex and multifactorial (61), public health messaging (alongside policy changes to ensure affordability) focusing on increasing fruit and vegetable intake as the one food group consistently associated with better health outcomes may be a sensible approach. A rapid review of the EWG has suggested a number of recommendations for better communication of the tool which, if adopted, may result in better adherence to the dietary guidelines (62).

There are some noted limitations of this analysis. The use of total fat as a diet quality measure is recognised to be crude and potentially misleading. We observed significant associations between meeting or getting higher scores on the fat component (i.e. eating, or being closer to eating, less than 35% of calories from fat) and both higher CAIDE score (greater risk for future dementia) and higher SBP, in the absence of any specific findings with SFA. Understanding the role of dietary fats in health has been a topic of much debate in the scientific literature and there is consensus that total fat content alone has little meaning for many health outcomes (63). Indeed, we know from many studies that nuts (source of omega 3 polyunsaturated fatty acids) and olive oil (source of monounsaturated fatty acids alongside some saturated and polyunsaturated fatty acids) is associated with favourable health outcomes (58, 64). This again suggests that further development of the EWG scores to better reflect the foods contributing to the macronutrients rather than the macronutrients themselves may be a more helpful approach to untangle the complexity of dietary fats. Finally, no participants met criteria for the sugars cut off applied to the dataset (≤5% calories from sugar), which is likely caused by the sugars calculated for PREVENT reflecting total sugars (glucose, galactose, fructose, sucrose, maltose and lactose) as opposed to free sugars (added sugars and naturally occurring sugars excluding galactose and lactose). Future nutritional analysis should consider a more detailed breakdown of sugars to better explore this component. The dietary data was collected from self-report questionnaires. Whilst the questionnaire has been validated in a number of settings (14), self-report of such data is known to be potentially fallible to bias through social desirability (65) and underreporting of energy intake (66). A further limitation to note is the lack of diversity of participants in the PREVENT cohort, with nearly all participants reporting their ethnicity as Caucasian (96.2%), and with an average of more than 16 years of education. This does not accurately reflect the UK and Irish populations, with a lack of ethnic diversity and a higher than average reported years of education. Finally, as this was a cross-sectional analysis, it is not possible to determine the directionality of any associations seen with reverse causality of poorer cardiovascular health driving dietary choices a possibility. Prospective studies are needed to confirm this relationship and replication of this analysis in the PREVENT cohort when the follow-up data is available will be important.

## Conclusions

This study developed scoring methodologies for a BEWG and GEWG score. Whilst there was no association between these scores and either risk for dementia or brain health in this mid-life cohort, there were significant associations between higher graded EWG scores and lower SBP, DBP and BMI. Adhering to fibre, fish, and fruit and vegetable were particularly associated with better cardiovascular health. Future research should further develop the EWG scores to reflect a food-based approach as opposed to the current reliance on macronutrient contributions to overall energy intake. Higher adherence to the EWG may be an important part of dementia risk reduction interventions through reductions in hypertension and obesity, both of which are important modifiable risk factors for dementia (49).

## Abbreviations

4MT: Four Mountains Test
AD: Alzheimer’s Disease *APOEε4* Apolipoprotein E Epsilon 4 BEWG Binary Eatwell Guide
BMI: Body Mass Index
CAIDE: Cardiovascular Risk Factors, Ageing and Dementia CI Confidence Interval
DBP: Diastolic Blood Pressure
EWG: Eatwell Guide
FDR: False Discovery Rate
FRS: Framingham Risk Score
GEWG: Graded Eatwell Guide
MEDAS: Mediterranean Diet Adherence Screener MedDiet Mediterranean Diet
MRI: Magnetic Resonance Imaging
NART: National Adult Reading Test
NS-SEC: National Statistics Socioeconomic Classification REC Research Ethics Committee
SBP: Systolic Blood Pressure
SCG-FFQ: Scottish Collaborative Group-Food Frequency Questionnaire
SD: Standard Deviation
SE: Standard Error
SES: Socioeconomic Status
SFA: Saturated Fatty Acids
UK: United Kingdom
UKWMCS UK: Women’s Cohort Study USA United States of Americ
WHR: Waist-to-Hip Ratio

## Data Availability

All data produced in the present study are available through the AD workbench provided by ADDI (https://www.alzheimersdata.org/ad-workbench)

## Acknowledgements

We would like to acknowledge the sites involved with the project, West London NHS Trust, NHS Lothian, Cambridgeshire and Peterborough NHS Foundation trust, Oxford Heath NHS Foundation trust and Trinity College. Special thanks also to the PREVENT participants, the participant panel, members of the Scientific Advisory Committee, and funders for their support of the PREVENT dementia programme.

## Authors’ contributions

SG, OS: Conceptualization, Methodology, Formal Analysis, Writing-Original Draft, Writing-Reviewing and Editing; AG, AJ, FCM, JM, RT, NW: Methodology, Formal Analysis, Writing-Reviewing and Editing; SP-S: Methodology, Writing-Reviewing and Editing; A-MM, GMT, CWR, ES: Supervision, Writing-Reviewing and Editing.

## Consent for publication

Not applicable.

## Availability of data and materials

Data from the PREVENT Dementia study can be accessed via the AD workbench (https://www.alzheimersdata.org/ad-workbench). DOI: https://doi.org/10.34688/PREVENTMAIN_BASELINE_700V1.

