## Supplementary Materials 1 for "Adherence to the Eatwell Guide and cardiometabolic, cognitive and neuroimaging parameters: An analysis from the PREVENT Dementia study"

|  | **BEWG Score** | | **GEWG Score** | |
| --- | --- | --- | --- | --- |
| **Carbohydrate** | 1 point: ≥50% of calories from carbohydrates  0 points: <50% calories from carbohydrates | | 5 points for ≥50% of calories from carbohydrates  4 points for ≥43.75% & <50% of calories from carbohydrates  3 points for ≥37.5% & <43.75% of calories from carbohydrates  2 points for ≥31.25% & <37.5% of calories from carbohydrates  1 point for ≥25% & <31.5% of calories from carbohydrates  0 points for <25% of calories from carbohydrates | |
| **Protein** | 1 point: ≥14.5% and ≤15.5% calories from protein  0 points: <14.5% or <15.5% calories from protein | | 5 points for ≥14.5% & ≤15.5% of calories from protein  4 points for ≥12.5% & <14.5% OR >15.5% & ≤17.5% calories from protein  3 points for ≥10.5% & <12.5% OR >17.5% & ≤19.5% calories from protein  2 points for ≥8.5% & <10.5% OR >19.5% & ≤21.5% calories from protein  1 point for ≥6.5% & <8.5% OR >21.5% & ≤23.5% calories from protein  0 points for <6.5% OR >23.5% calories from protein | |
| **Fat** | 1 point: ≤35% calories from fat  0 points: >35% calories from fat | | 5 points for ≤35% of calories from fat  4 points for >35% & ≤39.375% calories from fat  3 points for >39.375% & ≤43.75% calories from fat  2 points for >43.75% & ≤48.125% calories from fat  1 point for >48.125% & ≤52.5% calories from fat  0 points for >52.5% of calories from fat | |
| **SFA** | 1 point: ≤11% calories from SFA  0 points: >11% calories from SFA | | 5 points for ≤11% of calories from SFA  4 points for >11% & ≤12.375% calories from SFA  3 points for >12.375% & ≤13.75% calories from SFA  2 points for >13.75% & ≤15.125% calories from SFA  1 point for ≥15.125% & ≤16.5% calories from SFA  0 points for >16.5% of calories from SFA | |
| **Fibre** | 1 point: ≥22.6g/d  0 points: <22.6g/d | | 5 points for ≥22.6g/d  4 points for ≥19.775g/d & <22.6g/d  3 points for ≥16.95g/d & <19.775g/d  2 points for ≥14.125g/d & <16.95g/d  1 point for ≥11.3g/d & <14.125g/d  0 points for <11.3g/d | |
| **Sugars** | 1 point: ≤5% calories from sugar  0 points: >5% calories from sugar | | 5 points for ≤5% of calories from sugar  4 points for >5 & ≤5.625 of calories from sugar  3 points for >5.625 & ≤6.25 of calories from sugar  2 points for >6.25 & ≤6.875 of calories from sugar  1 point for >6.875 & ≤7.5 of calories from sugar  0 points for >7.5% of calories from sugar | |
| **Salt (Sodium)** | 1 point: ≤2363mg/d  0 points: >2363mg/d | | 5 points for ≤2363mg/d  4 points for >2363mg/d & ≤2658.375mg/d  3 points for >2658.375mg/d & ≤2953.65mg/d  2 points for >2953.65mg/d & ≤3249.025mg/d  1 point for >3249.025mg/d & ≤3544.5mg/d  0 points for >3544.5mg/d | |
| **Total kilocalories** | Male  1 point: ≥2250kcal and ≤2750kcal  0 points: <2250kcal or >2750kcal | Female  1 point: ≥1800kcal and ≤2200kcal  0 points: <1800kcal or >2200kcal | Male  5 points for <2750kcal  4 points for ≥2750kcal & <3000kcal  3 points for ≥3000kcal & <3250kcal  2 points for ≥3250kcal & <3500kcal  1 point for ≥3500kcal & <3750kcal  0 points for <3750kcal | Female  5 points for <2200kcal  4 points for ≥2200kcal & <2400kcal  3 points for ≥2400kcal & <2600kcal  2 points for ≥2600kcal & <2800kcal  1 point for ≥2800kcal & <3000kcal  0 points for <3000kcal |
| **Fruit and vegetables** | 1 point: ≥400g/d  0 points: <400g/d | | 5 points for ≥400g/d  4 points for ≥350g/d & <400g/d  3 points for ≥300g/d & <350g/d  2 points for ≥250g/d & <300g/d  1 point for ≥200g/d & <250g/d  0 points for <200g/d | |
| **Fish** | 1 point: ≥20g/d  0.5 points: no oily fish consumed  0 points: <20g/d | | Total fish  5 points for ≥20g/d  4 points for ≥17.5g/d & <20g/d  3 points for ≥15g/d & <17.5g/d  2 points for ≥12.5g/d & <15g/d  1 point for ≥10g/d & <12.5g/d  0 points for <10g/d | Oily fish  5 points for ≥10g/d  4 points for ≥8.75g/d & <10g/d  3 points for ≥7.5g/d & <8.75g/d  2 points for ≥6.25g/d & <7.5g/d  1 point for ≥5g/d & <6.25g/d  0 points for <5g/d |
|  |  |  | Total component value = mean of total and oily fish scores | |
| **Red and processed meat** | 1 point: ≤70g/d  0 points: >70g/d | | 5 points for ≤70g/d  4 points for ≥70g/d & <78.5g/d  3 points for ≥78.5g/d & <87.5g/d  2 points for ≥87.5g/d & <96.25g/d  1 point for ≥96.25g/d & <105g/d  0 points for ≥105g/d | |
| **Water** | 1 point: ≥6 portions/d  0 points: <6 portions/d | | 5 points for ≥6 portions/d  4 points for ≥5.25 portions/d & <6 portions/d  3 points for ≥4.5 portions/d & <5.25 portions/d  2 points for ≥3.75 portions/d & <4.5 portions/d  1 point for ≥3 portions/d & <3.75 portions/d  0 points for <3 portions/d | |
| **Total score** | Sum of individual components | | Sum of individual components | |

Supplementary Table S1: Scoring methodologies for BEWG score and GEWG graded score. d: day; BEWG: Binary Eatwell Guide; g: grams; GEWG: Graded Eatwell Guide; SFA: saturated fatty acids

| **Score component** | **Score allocation** |
| --- | --- |
| Age | < 47 years: 0 points  47-53 years: 3 points  > 53 years: 5 points |
| Education | ≥ 10 years: 0 points  7-9 years: 3 points  0-6 years: 6 points |
| Sex | Women: 0 points  Men: 1 point |
| SBP | ≤ 140 mmHg: 0 points  > 140 mmHg: 2 points |
| BMI | ≤ 30 kg/m^2^: 0 points  > 30 kg/m^2^: 2 points |
| Total Cholesterol | ≤ 6.5 mmol/L: 0 points  > 6.5 mmol/L: 1 point |
| Physical Activity | Active: 0 points  Inactive: 1 point |
| *APOEε4* carrier status | Non-carrier: 0 points  Carrier: 2 points |

Supplementary Table S2: Table of scoring components and weight used to create the CAIDE score. BMI: body mass index; CAIDE: Cardiovascular Risk Factors Ageing and Dementia; L: litre; m: metres; mmHG: millimetres of mercury; mmol: millimolar; kg: kilograms SBP: systolic blood pressure.

| **Score component** | **FRS** | **QRisk3** |
| --- | --- | --- |
| Age | **✓** | **✓** |
| Anti-hypertensive use | **✓** | **✓** |
| Diabetes | **✓** | **✓** |
| SBP | **✓** | **✓** |
| Sex | **✓** | **✓** |
| Smoking | **✓** | **✓** |
| Atrial fibrillation |  | **✓** |
| Atypical antipsychotics |  | **✓** |
| Cholesterol to HDL ratio |  | **✓** |
| Erectile dysfunction |  | **✓** |
| Ethnicity |  | **✓** |
| Height |  | **✓** |
| Kidney disease |  | **✓** |
| Migraine |  | **✓** |
| Rheumatoid arthritis |  | **✓** |
| Severe mental illness |  | **✓** |
| Standard deviation of SBP |  | **✓** |
| Steroids |  | **✓** |
| Systemic lupus erythematosus |  | **✓** |
| Weight |  | **✓** |

*Supplementary Table S3: Table of scoring components used to create the FRS and QRisk3 scores. BMI: body mass; FRS: Framingham Risk Score; HDL: high density lipoprotein; SBP: systolic blood pressure.*

| **Score component** | **BEWG**  **n (%)** | **GEWG**  **mean (SD)** |
| --- | --- | --- |
| Carbohydrate | 70 (13.5) | 3.44 (1.00) |
| Protein | 93 (18.0) | 3.68 (1.00) |
| Fat | 199 (38.5) | 4.01 (1.02) |
| SFA | 75 (14.5) | 2.48 (1.67) |
| Fibre | 116 (22.4) | 2.54 (1.78) |
| Sugars | 0 (0) | 0.00 (0.04) |
| Salt | 241 (46.6) | 3.38 (1.90) |
| Total kilocalories | 116 (22.4) | 4.34 (1.40) |
| Fruit and vegetables | 250 (48.4) | 3.75 (1.45) |
| Fish | 375 (75.9) | 3.83 (1.76) |
| Red and processed meat | 415 (80.3) | 4.41 (1.38) |
| Water | 307 (59.4) | 4.02 (1.48) |

*Supplementary Table S4: Breakdown of contributing components to the BEWG and GEWG graded scores. BEWG: Binary Eatwell Guide; GEWG: Graded Eatwell Guide; SD: standard deviation; SFA: saturated fatty acids.*

| **BEWG Component** |  | **Unadjusted model** | | |  | **Fully adjusted model** | | |
| --- | --- | --- | --- | --- | --- | --- | --- | --- |
|  | **β** | **SE** | **95% CI** | **p** | **β** | **SE** | **95% CI** | **p** |
| **Carbohydrate** | 0.12 | 0.36 | -0.60, 0.85 | 0.73 | 0.18 | 0.35 | -0.60, 0.85 | 0.61 |
| **Protein** | -0.44 | 0.32 | -1.08, 0.21 | 0.18 | -0.15 | 0.32 | -0.79, 0.48 | 0.63 |
| **Fat** | 0.55 | 0.25 | 0.04, 1.06 | 0.03 | 0.61 | 0.25 | 0.12, 1.11 | 0.01 |
| **SFA** | -0.004 | 0.35 | -0.71, 0.70 | 0.99 | 0.01 | 0.34 | -0.67, 0.70 | 0.97 |
| **Fibre** | 0.12 | 0.30 | -0.48, 0.72 | 0.69 | 0.16 | 0.29 | -0.43, 0.74 | 0.60 |
| **Salt** | 0.23 | 0.25 | -0.26, 0.73 | 0.35 | 0.24 | 0.24 | -0.25, 0.72 | 0.33 |
| **Total kilocalories** | 0.05 | 0.30 | -0.54, 0.65 | 0.86 | -0.04 | 0.29 | -0.62, 0.54 | 0.89 |
| **Fruit and vegetables** | -0.34 | 0.25 | -0.83, 0.16 | 0.18 | -0.30 | 0.24 | -0.78, 0.19 | 0.22 |
| **Fish** | 0.18 | 0.30 | -0.36, 0.82 | 0.54 | 0.28 | 0.29 | -0.24, 0.91 | 0.33 |
| **Red and processed meat** | 0.25 | 0.31 | -0.37, 0.88 | 0.42 | 0.31 | 0.30 | -0.29, 0.92 | 0.30 |
| **Water** | -0.29 | 0.25 | -0.79, 0.22 | 0.26 | -0.26 | 0.24 | -0.75, 0.23 | 0.28 |

*Supplementary Table S5: Generalised additive models for each component of the BEWG score with the CAIDE score as the outcome. Fully adjusted model includes parental history of dementia, physical activity score and SES group as covariates. BEWG: Binary Eatwell guide; CI: confidence interval; SES: socioeconomic status; SFA: saturated fatty acids.*

| **GEWG Component** |  | **Unadjusted model** | | |  | **Fully adjusted model** | | |
| --- | --- | --- | --- | --- | --- | --- | --- | --- |
|  | **β** | **SE** | **95% CI** | **p** | **β** | **SE** | **95% CI** | **p** |
| **Carbohydrate** | 0.003 | 0.12 | -0.25, 0.25 | 0.98 | 0.04 | 0.12 | -0.20, 0.28 | 0.73 |
| **Protein** | -0.08 | 0.12 | -0.33, 0.17 | 0.53 | 0.04 | 0.12 | -0.20, 0.28 | 0.75 |
| **Fat** | 0.19 | 0.12 | -0.06, 0.43 | 0.12 | 0.24 | 0.12 | 0.01, 0.48 | 0.04 |
| **SFA** | 0.06 | 0.07 | -0.09, 0.21 | 0.40 | 0.09 | 0.07 | -0.06, 0.23 | 0.24 |
| **Fibre** | -0.06 | 0.07 | -0.20, 0.08 | 0.43 | -0.05 | 0.07 | -0.19, 0.08 | 0.44 |
| **Sugars** | 0.05 | 2.83 | -5.62, 5.72 | 0.99 | -0.31 | 2.75 | -5.81, 5.19 | 0.91 |
| **Salt** | 0.02 | 0.07 | -0.11, 0.15 | 0.72 | 0.03 | 0.06 | -0.09, 0.16 | 0.60 |
| **Total kilocalories** | 0.05 | 0.09 | -0.13, 0.23 | 0.57 | 0.10 | 0.09 | -0.07, 0.28 | 0.24 |
| **Fruit and vegetables** | -0.14 | 0.09 | -0.31, 0.03 | 0.11 | -0.09 | 0.08 | -0.26, 0.07 | 0.26 |
| **Fish** | 0.07 | 0.07 | -0.07, 0.21 | 0.31 | 0.09 | 0.07 | -0.05, 0.23 | 0.18 |
| **Red and processed meat** | 0.02 | 0.09 | -0.16, 0.20 | 0.78 | 0.05 | 0.09 | -0.13, 0.22 | 0.57 |
| **Water** | -0.11 | 0.08 | -0.28, 0.05 | 0.18 | -0.10 | 0.08 | -0.27, 0.06 | 0.20 |

*Supplementary Table S6: Generalised additive models for each component of the GEWG score with the CAIDE score as the outcome. Fully adjusted model includes parental history of dementia, physical activity score and SES group as covariates. CI: confidence interval; GEWG: Graded Eatwell guide; SES: socioeconomic status; SFA: saturated fatty acids.*

| **GEWG Component** | **Unadjusted model** | | | | **Fully adjusted model** | | | |
| --- | --- | --- | --- | --- | --- | --- | --- | --- |
|  | **β** | **SE** | **95% CI** | **p** | **β** | **SE** | **95% CI** | **p** |
| Carbohydrate | 0.55 | 0.68 | -0.82, 1.92 | 0.42 | 0.61 | 0.64 | -0.66, 1.88 | 0.34 |
| Protein | 0.34 | 0.68 | -1.03, 1.71 | 0.62 | -0.40 | 0.65 | -1.69, 0.89 | 0.54 |
| Fat | 1.67 | 0.66 | 0.34, 3.00 | 0.01 | 1.53 | 0.63 | 0.28, 2.78 | 0.01 |
| SFA | -0.37 | 0.41 | -1.19, 0.45 | 0.37 | -0.19 | 0.38 | -0.96, 0.58 | 0.62 |
| Fibre | -0.84 | 0.38 | -1.60, -0.07 | 0.03 | -0.97 | 1.30 | -1.69, -0.26 | 0.007 |
| Sugars | 2.76 | 15.57 | -28.38, 33.91 | 0.86 | -4.96 | 14.49 | -33.93, 24.01 | 0.73 |
| Salt | -0.02 | 0.36 | -0.74, 0.71 | 0.97 | 0.35 | 0.34 | -0.33, 1.02 | 0.31 |
| Total kilocalories | 0.67 | 0.49 | -0.31, 1.65 | 0.17 | 0.32 | 0.46 | -0.60, 1.24 | 0.49 |
| Fruit and vegetables | -1.48 | 0.47 | -2.41, -0.55 | 0.002 | -1.09 | 0.44 | -1.97, -0.20 | 0.01 |
| Fish | -1.01 | 0.39 | -1.78, -0.23 | 0.01 | -1.03 | 0.36 | -1.75, -0.30 | 0.005 |
| Red and processed meat | -1.08 | 0.49 | -2.07, -0.10 | 0.03 | -0.90 | 0.46 | -1.82, 0.03 | 0.05 |
| Water | -1.02 | 0.46 | -1.94, -0.10 | 0.03 | -0.70 | 0.43 | -1.55, 0.16 | 0.10 |

*Supplementary Table S7: Generalised additive models for each component of the GEWG score with systolic blood pressure as the outcome. Fully adjusted model includes age, sex, education, APOEε4, parental history of dementia, physical activity score and SES group as covariates. CI: confidence interval; GEWG: Graded Eatwell guide; SES: socioeconomic status; SFA: saturated fatty acids.*

| **GEWG Component** | **Unadjusted model** | | | | **Fully adjusted model** | | | |
| --- | --- | --- | --- | --- | --- | --- | --- | --- |
|  | **β** | **SE** | **95% CI** | **p** | **β** | **SE** | **95% CI** | **p** |
| Carbohydrate | -0.47 | 0.42 | -1.31, 0.37 | 0.27 | -0.44 | 0.39 | -1.23, 0.35 | 0.27 |
| Protein | 0.13 | 0.42 | -0.71, 0.98 | 0.75 | -0.31 | 0.40 | -1.11, 0.50 | 0.45 |
| Fat | 0.69 | 0.41 | -0.13, 1.52 | 0.09 | 0.61 | 0.39 | -0.17, 1.39 | 0.12 |
| SFA | -0.22 | 0.25 | -0.73, 0.28 | 0.38 | -0.10 | 0.24 | -0.58, 0.37 | 0.66 |
| Fibre | -0.57 | 0.24 | -1.05, -0.10 | 0.02 | -0.61 | 0.22 | -1.05, -0.16 | 0.006 |
| Sugars | -5.60 | 9.59 | -24.77, 13.58 | 0.56 | -11.93 | 8.99 | -29.91, 6.05 | 0.19 |
| Salt | 0.05 | 0.22 | -0.40, 0.49 | 0.84 | 0.24 | 0.21 | -0.19, 0.66 | 0.26 |
| Total kilocalories | 0.43 | 0.30 | -0.17, 1.03 | 0.16 | 0.19 | 0.29 | -0.39, 0.76 | 0.51 |
| Fruit and vegetables | -1.08 | 0.29 | -1.65, -0.51 | 0.0002 | -0.79 | 0.27 | -1.34, -0.24 | 0.004 |
| Fish | -0.41 | 0.24 | -0.89, 0.07 | 0.09 | -0.39 | 0.23 | -0.84, 0.06 | 0.08 |
| Red and processed meat | -0.92 | 0.30 | -1.52, -0.31 | 0.003 | -0.77 | 0.29 | -1.34, -0.20 | 0.007 |
| Water | -0.28 | 0.28 | -0.95, 0.19 | 0.18 | -0.19 | 0.27 | -0.72, 0.34 | 0.47 |

*Supplementary Table S8: Generalised additive models for each component of the GEWG score with diastolic blood pressure as the outcome. Fully adjusted model includes age, sex, education, APOEε4, parental history of dementia, physical activity score and SES group as covariates. CI: confidence interval; GEWG: Graded Eatwell guide; SES: socioeconomic status; SFA: saturated fatty acids.*

| **GEWG Component** | **Unadjusted model** | | | | **Fully adjusted model** | | | |
| --- | --- | --- | --- | --- | --- | --- | --- | --- |
|  | **β** | **SE** | **95% CI** | **p** | **β** | **SE** | **95% CI** | **p** |
| Carbohydrate | -0.09 | 0.23 | -0.55, 0.37 | 0.70 | -0.08 | 0.22 | -0.53, 0.36 | 0.71 |
| Protein | -0.45 | 0.23 | -0.91, 0.002 | 0.047 | -0.40 | 0.22 | -0.85, 0.05 | 0.07 |
| Fat | 0.13 | 0.22 | -0.32, 0.58 | 0.56 | 0.15 | 0.22 | -0.29, 0.59 | 0.50 |
| SFA | -0.19 | 0.14 | -0.46, 0.09 | 0.18 | -0.13 | 0.13 | -0.40, 0.14 | 0.33 |
| Fibre | -0.10 | 0.13 | -0.35, 0.16 | 0.46 | -0.04 | 0.13 | -0.30, 0.21 | 0.72 |
| Sugars | -4.89 | 5.19 | -15.28, 5.49 | 0.35 | -6.72 | 5.04 | -16.80, 3.37 | 0.18 |
| Salt | -0.26 | 0.12 | -0.50, -0.02 | 0.03 | -0.23 | 0.12 | -0.46, 0.009 | 0.05 |
| Total kilocalories | -0.32 | 0.16 | -0.64, 0.009 | 0.05 | -0.30 | 0.16 | -0.62, 0.02 | 0.06 |
| Fruit and vegetables | -0.54 | 0.16 | -0.85, -0.23 | 0.0006 | -0.38 | 0.15 | -0.69, -0.07 | 0.01 |
| Fish | -0.03 | 0.13 | -0.29, 0.23 | 0.79 | 0.03 | 0.13 | -0.22, 0.29 | 0.81 |
| Red and processed meat | -0.42 | 0.16 | -0.74, -0.09 | 0.01 | -0.31 | 0.16 | -0.63, 0.01 | 0.06 |
| Water | 0.12 | 0.15 | -0.19, 0.42 | 0.46 | 0.17 | 0.15 | -0.13, 0.47 | 0.26 |

*Supplementary Table S8: Generalised additive models for each component of the GEWG score with BMI as the outcome. Fully adjusted model includes age, sex, education, APOEε4, parental history of dementia, physical activity score and SES group as covariates. BMI: body mass index; CI: confidence interval; GEWG: Graded Eatwell guide; SES: socioeconomic status; SFA: saturated fatty acids.*

| **Outcome** | **Low** | | | | **Middle** | | | | **High** | | | | **Not in employment** | | | |
| --- | --- | --- | --- | --- | --- | --- | --- | --- | --- | --- | --- | --- | --- | --- | --- | --- |
|  | **β** | **SE** | **95% CI** | **p** | **β** | **SE** | **95% CI** | **p** | **β** | **SE** | **95% CI** | **p** | **β** | **SE** | **95% CI** | **p** |
| **CAIDE^1^** | 0.02 | 0.09 | -0.16, 0.19 | 0.86 | 0.01 | 0.05 | -0.09, 0.12 | 0.78 | 0.02 | 0.02 | -0.03, 0.07 | 0.39 | -0.03 | 0.05 | -0.13, 0.08 | 0.59 |
| **SBP^1^** | 0.19 | 0.38 | -0.56, 0.95 | 0.61 | -0.33 | 0.29 | -0.92, 0.26 | 0.27 | -0.25 | 0.13 | -0.51, 0.01 | 0.06 | -0.10 | 0.31 | -0.73, 0.53 | 0.75 |
| **DBP^1^** | 0.11 | 0.20 | -0.29, 0.52 | 0.58 | -0.20 | 0.18 | -0.64, 0.06 | 0.10 | -0.14 | 0.08 | -0.30, 0.03 | 0.10 | -0.13 | 0.21 | -0.55, 0.30 | 0.56 |
| **BMI^1^** | 0.12 | 0.12 | -0.12, 0.36 | 0.34 | -0.16 | 0.10 | -0.36, 0.04 | 0.11 | -0.09 | 0.05 | -0.18, 0.007 | 0.07 | 0.02 | 0.11 | -0.19, 0.23 | 0.85 |
| **WHR^1^** | -0.003 | 0.003 | -0.01, 0.005 | 0.47 | 0.0002 | 0.001 | -0.002, 0.003 | 0.99 | -0.0004 | 0.0009 | -0.002, 0.001 | 0.67 | 0.0002 | 0.001 | -0.003, 0.003 | 0.90 |
| **FRS^2^** | -0.07 | 0.23 | -0.53, 0.38 | 0.75 | -0.14 | 0.13 | -0.40, 0.13 | 0.31 | -0.03 | 0.05 | -0.14, 0.08 | 0.55 | -0.13 | 0.26 | -0.44, 0.19 | 0.42 |
| **QRisk3^2^** | 0.009 | 0.12 | -0.23, 0.25 | 0.94 | -0.07 | 0.07 | -0.22, 0.08 | 0.37 | 0.01 | 0.04 | -0.06, 0.09 | 0.71 | -0.06 | 0.09 | -0.23, 0.12 | 0.53 |
| **4MT^3^** | 0.07 | 0.12 | -0.18, 0.32 | 0.58 | 0.06 | 0.09 | -0.12, 0.24 | 0.51 | 0.002 | 0.04 | -0.07, 0.08 | 0.96 | 0.10 | 0.11 | -0.11, 0.32 | 0.34 |
| **WMLV^4^** | -0.01 | 0.01 | -0.04, 0.01 | 0.40 | 0.009 | 0.006 | -0.003, 0.02 | 0.14 | -0.003 | 0.003 | -0.01, 0.004 | 0.36 | -0.0007 | 0.007 | -0.01, 0.01 | 0.92 |
| **Left hippocampus^4^** | -11.18 | 8.81 | -28.80, -6.43 | 0.22 | 6.19 | 6.77 | -7.35, 19.72 | 0.36 | -7.81 | 3.04 | -13.90, -1.73 | 0.01 | 4.93 | 6.84 | -8.74, 18.61 | 0.47 |
| **Right hippocampus^4^** | -2.35 | 10.80 | -21.84, 21.37 | 0.98 | 9.08 | 7.64 | -6.21, 24.36 | 0.24 | -4.56 | 3.37 | -11.29, 2.18 | 0.18 | 3.99 | 7.83 | -11.67, 19.66 | 0.61 |
| **Left hippocampal thickness^4^** | 0.002 | 0.002 | -0.001, 0.006 | 0.23 | -0.001 | 0.002 | -0.005, 0.002 | 0.47 | 0.0009 | 0.0006 | -0.0003, 0.002 | 0.14 | 0.003 | 0.002 | -0.001, 0.007 | 0.14 |
| **Right hippocampal thickness^4^** | 0.003 | 0.002 | -0.001, 0.007 | 0.19 | -0.002 | 0.002 | -0.005, 0.001 | 0.29 | 0.0008 | 0.0006 | -0.0004, 0.002 | 0.21 | 0.002 | 0.002 | -0.001, 0.006 | 0.24 |
| **Self-reported healthy eating^5^** | 0.03 | 0.06 | -0.08, 0.15 | 0.59 | 0.09 | 0.05 | -0.0006, 0.19 | 0.06 | 0.11 | 0.03 | 0.05, 0.17 | <0.001 | 0.20 | 0.11 | 0.008, 0.47 | 0.07 |

*Supplementary Table S9: Fully adjusted generalised additive models for the GEWG score with outcome measures of interest by SES group. ^1^Low n=40, middle n=81, high n=334, not in employment n=62, covariates parental history of dementia and physical activity scores for CAIDE analysis, covariates age, sex, years of education, APOEε4, parental history of dementia and physical activity scores for SBP. DBP, BMI and WHR analysis; ^2^Low n=40, middle n=77, high n=325, not in employment n=61, covariates years of education, APOEε4, parental history of dementia and physical activity score; ^3^Low n=25, middle n=49, high n=230, not in employment n=38, covariates age, sex, years of education, APOEε4, parental history of dementia, NART score and physical activity score; ^4^Low n=39, middle n=81, high n=332, not in employment n=62, covariates age, sex, years of education, APOEε4, parental history of dementia, physical activity score and intracranial volume, WMLV cube transformed; ^5^Low n=40, middle n=81, high n=333, not in employment n=62; covariates age, sex, years of education, APOEε4, parental history of dementia, physical activity score. BMI: body mass index; CI: confidence interval; GEWG: Graded Eatwell guide; FRS: Framingham Risk Score; SES: socioeconomic status; SFA: saturated fatty acids; WHR: waist-to-hip ratio; WMLV: white matter lesion volume.*

| **CAIDE score and cardiovascular outcome variables** | **EWG Low Adherence (n=161)** | **EWG High Adherence (n=356)** |
| --- | --- | --- |
| CAIDE score (mean, SD) | 5.86 (3.08) | 5.99 (2.71) |
| SBP (mmHg) (mean, SD) | 127.24 (15.48) | 123.85 (15.48) |
| DBP (mmHg) (mean, SD) | 77.56 (9.74) | 75.66 (9.45) |
| BMI (kg/m^2^) (mean, SD) | 27.23 (5.11) | 27.22 (5.23) |
| WHR (mean, SD) | 0.89 (0.11) | 0.87 (0.10) |
| **Cardiovascular risk scores** | **EWG Low Adherence (n=156)** | **EWG High Adherence (n=347)** |
| FRS (mean, SD) | 9.37 (7.02) | 8.39 (6.10) |
| QRisk3 (mean, SD) | 5.03 (4.65) | 4.71 (3.74) |
| **Cognition** | **EWG Low Adherence (n=119)** | **EWG High Adherence (n=223)** |
| 4MT total score (mean, SD) | 9.67 (3.67) | 9.95 (3.24) |
| **Neuroimaging** | **EWG Low Adherence (n=147)** | **EWG High Adherence (n=332)** |
| White matter lesion volume (mL) (mean, SD) | 1.21 (0.40) | 1.18 (0.36) |
| Left hippocampus volume (mm^3^) (mean, SD) | 4076.11 (365.86) | 4018.00 (389.43) |
| Right hippocampus volume (mm^3^) (mean, SD) | 4190.70 (418.36) | 4145.50 (429.43) |
| Left hippocampus thickness (mm) (mean, SD) | 2.44 (0.07) | 2.44 (0.07) |
| Right hippocampus thickness (mm) (mean, SD) | 2.43 (0.07) | 2.43 (0.06) |

*Supplementary Table S10: Demographic and descriptive statistics of sample included in Eatwell Guide score analysis spit by the median EWG score. 4MT: Four Mountains Test; BMI: body mass index; DBP: diastolic blood pressure; EWG: Eatwell Guide; FRS: Framingham Risk Score; mL: millilitres; mm: millimetres; SBP: systolic blood pressure; SD: standard deviation; SES: socioeconomic status.*

| **Dietary score** | Unadjusted | | | | Fully adjusted | | | |
| --- | --- | --- | --- | --- | --- | --- | --- | --- |
|  | β | **SE** | **95% CI** | **p** | β | **SE** | **95% CI** | **p** |
| **CAIDE^1^** | | | | | | | | |
| **EWG (Low)** | -0.13 | 0.27 | -0.66, 0.39 | 0.62 | -0.16 | 0.26 | -0.67, 0.35 | 0.54 |
| **Systolic Blood Pressure^1^** | | | | | | | | |
| **EWG (Low)** | 3.38 | 1.47 | 0.50, 6.26 | 0.02 | 3.38 | 1.49 | 0.46, 6.30 | 0.02 |
| **Diastolic Blood Pressure^1^** | | | | | | | | |
| **EWG (Low)** | 1.89 | 0.91 | 0.12, 3.67 | 0.04 | 1.91 | 0.92 | 0.11, 3.71 | 0.04 |
| **BMI^1^** | | | | | | | | |
| **EWG (Low)** | 0.01 | 0.49 | -0.95, 0.98 | 0.98 | 0.04 | 0.49 | -0.92, 0.99 | 0.94 |
| **WHR^1^** | | | | | | | | |
| **EWG (Low)** | 0.01 | 0.01 | -0.006, 0.03 | 0.18 | 0.01 | 0.01 | -0.007, 0.03 | 0.20 |
| **FRS^2^** | | | | | | | | |
| **EWG (Low)** | 0.98 | 0.62 | -0.23, 2.19 | 0.11 | 0.97 | 0.62 | -0.24, 2.19 | 0.12 |
| **QRisk3^2^** | | | | | | | | |
| **EWG (Low)** | 0.32 | 0.39 | -0.44, 1.09 | 0.41 | 0.31 | 0.39 | -0.46, 1.08 | 0.43 |
| **4MT Total Score^3^** | | | | | | | | |
| **EWG (Low)** | 0.98 | 0.52 | -0.05, 2.00 | 0.06 | 1.14 | 0.53 | 0.10, 2.17 | 0.03 |
| **White Matter Lesion Volume^4^** | | | | | | | | |
| **EWG (Low)** | 0.03 | 0.04 | -0.04, 0.10 | 0.43 | 0.04 | 0.04 | -0.04, 0.11 | 0.33 |
| **Left Hippocampus^4^** | | | | | | | | |
| **EWG (Low)** | 53.84 | 30.27 | -5.48, 113.16 | 0.08 | 57.10 | 30.70 | -3.07, 117.26 | 0.06 |
| **Right Hippocampus^4^** | | | | | | | | |
| **EWG (Low)** | 40.48 | 33.86 | -25.87, 106.84 | 0.23 | 46.19 | 34.27 | -20.98, 113.37 | 0.18 |
| **Left Hippocampal Thickness^4^** | | | | | | | | |
| **EWG (Low)** | -0.003 | 0.007 | -0.02, 0.01 | 0.65 | -0.001 | 0.007 | -0.01, 0.01 | 0.87 |
| **Right Hippocampal Thickness^4^** | | | | | | | | |
| **EWG (Low)** | -0.002 | 0.007 | -0.01, 0.01 | 0.75 | -0.0002 | 0.007 | -0.01, 0.01 | 0.98 |

*Supplementary Table S11: Unadjusted and fully adjusted logistic regression models for the EWG score split into low and high adherence by the median. ^1^n=517, ^2^n=503, ^3^n=342, ^4^n=479. Covariates parental history of dementia and physical activity scores for CAIDE analysis, covariates age, sex, years of education, APOEε4, parental history of dementia and physical activity scores for SBP. DBP, BMI and WHR analysis; covariates age, sex, years of education, APOEε4, parental history of dementia, physical activity scores and estimated intracranial volume for left nd right hippocampal volume and left and right hippocampal thickness analysis. BMI: body mass index; CI: confidence interval; GEWG: Graded Eatwell guide; FRS: Framingham Risk Score; SES: socioeconomic status; SFA: saturated fatty acids; WHR: waist-to-hip ratio; WMLV: white matter lesion volume.*
